# The Role of CRP, Interleukin-6 and Their Derived Immune-Inflammatory Indices in Early Prediction of Severity and Mortality of COVID-19 Patients

**DOI:** 10.1101/2021.06.28.21259644

**Authors:** Sara I. Taha, Aalaa K. Shata, Eman M. El-Sehsah, Shaimaa H. Fouad, Aya H. Moussa, Shaimaa A. Abdalgeleel, Nouran M. Moustafa, Mariam K. Youssef

## Abstract

**Background:** In coronavirus disease 2019 (COVID-19), finding sensitive biomarkers is critical for detecting severe cases early and intervening effectively.

**Objectives:** To compare and evaluate the predictive value of C-reactive protein (CRP), interleukin-6 (IL-6), and their derived immune-inflammatory indices (CRP/albumin (CRP/alb), lymphocyte/CRP (L/CRP), and lymphocyte/IL-6 (L/IL-6)) in COVID-19 patients.

**Methods:** On admission, 85 confirmed COVID-19 patients, their measured and collected laboratory data were obtained and compared.

**Results:** Levels of CRP, IL-6, and CRP/alb were significantly higher (P=0.001) in severe patients and non-survivors, but L/CRP and L/IL-6 were significantly lower (P=0.001). We observed the best predictive performance for COVID-19 severity at 1.65 for CRP/alb and 260.86 for L/CRP with 84.7% diagnostic accuracy for both. The best diagnostic accuracy for COVID-19 in-hospital mortality was 87.1% by IL-6 at 120 pg/ml and 85.9% by L/IL-6 at 5.40. The multi-marker prediction surpassed the performance of a single biomarker prediction. IL-6 was an independent risk factor associated with severe disease development (odds ratio (OR): 1.033; 95% confidence interval (CI): 1.002-1.066).

**Conclusions:** Pretreatment values of CRP, IL-6, and their derived indices could be included in the diagnostic work-up of COVID-19 to determine the severity and predict the outcome.

## Introduction

The pandemic of coronavirus disease 2019 (COVID-19) is presently sweeping the globe, affecting millions of individuals.^**1**^ COVID-19 is more contagious than seasonal influenza, has a longer incubation time, and is linked to a greater hospitalization rate and overall mortality. The virus spreads through the respiratory tract and quickly progresses to severe infection, multi-organ failure, and even death.^**2**^

Although most COVID-19 patients recover quickly, patients with moderate severity, particularly those with risk factors like old age, obesity, and associated co-morbidities, can rapidly worsen and become severe, increasing ICU admission and mechanical ventilation, indicating a high mortality rate.^**3**^ Thus, early identification of potentially severe patients is critical for halting disease progression at an early stage.

COVID-19 progression may be facilitated by an abnormal inflammatory response and cytokine storm.^**4**^ Therefore, this study was performed to evaluate the predictive efficacy of individual and combined baseline inflammatory indicators; C-reactive protein (CRP) and interleukin-6 (IL-6) together with their derived immune-inflammatory indices (CRP/albumin (CRP/alb), lymphocyte/CRP (L/CRP), and lymphocyte/IL-6 (L/IL-6)) in a cohort of Egyptian COVID-19 patients.

## Materials & methods

### Patients

This cohort observational study recruited 85 adult patients (aged ≥ 18 years) from COVID-19 Isolation Hospitals of Ain-Shams University, Cairo, Egypt. A real-time polymerase chain reaction (RT-PCR) test confirmed COVID-19 in all of the patients. We excluded pregnant women and patients with any hematological diseases, immunodeficiencies, or autoimmune disorders. All participants were classified according to their disease severity using the COVID-19 management guideline provided by Ain Shams University Hospitals.^**5**^

### Data collection

From medical records, epidemiological and clinical data (such as age, sex, underlying diseases or comorbidities, length of stay in the hospital, the requirement for ICU admission, and patients’ outcomes) were collected, as well as laboratory findings from the same day (such as complete blood differential counts and CRP).

### Blood sampling and analysis

Venous blood samples were taken via venipuncture from all enrolled patients into biochemistry tubes covered with gel for serum separation. The samples were allowed to clot for 30 mins before being centrifuged for 10 mins at 2,500 g. The serum was immediately used to determine albumin level using a Beckman Coulter AU480 autoanalyzer (Beckman Coulter, Inc., USA). The dynamic range of albumin was 1.5-6.0 g/dL. The remaining sera were kept at -80°C until they were tested for IL-6 using a commercially available Human IL-6 ELISA Kit from RayBiotech Life, Inc., GA, USA [CODE: ELH-IL6-1]. According to the supplier’s instructions, the assay was carried out with a detection range of 3 pg/ml - 1000 pg/ml.

### Calculations

Derived immune-inflammatory indices were calculated as follows: L/CRP = lymphocyte (number/μl)/CRP (mg/dl); L/IL-6 = lymphocyte (number/μl)/IL-6 (pg/ml); CRP/alb = CRP (mg/dl)/albumin (g/dl).

### Statistical analysis

Statistical analysis was performed using the SPSS 20.0. Descriptive statistics were done using median and percentiles for quantitative nonparametric measures, while categorized data were described in numbers and percentages. The Wilcoxon Rank Sum test and Chi-square test were used to compare two groups of quantitative nonparametric and qualitative data, respectively. The receiver operating characteristic (ROC) curve assessed the predictive performance of each pretreatment biomarker alone and the clinical benefit of the biomarkers combination. The odds ratio (OR) and 95% confidence interval (CI) were calculated using logistic regression analysis. The significant probability of error was set at 0.05.

## Results

### Demographic and Clinical Characterization of the Study Cohort

Of the 85 included patients, 48 (56.5%) were males, and 37 (43.5%) were females with a median (IQR) age of 55 years (42 – 65). Of all, 54.1% (n=46) were non-severe while 45.9% (n=39) were severe. The most prevalent presenting symptoms were dyspnea (51.8%) and fever (40%). Hypertension (40.0%) and diabetes mellitus (36.5%) were the most common associated comorbidities. The median (IQR) duration of hospitalization was 10 days (6 – 17), during which 38.8% (n=33) of patients required ICU admission, and 24.7% (n=21) died. Baseline findings of the studied population (n=85) are demonstrated in **table 1**.

**Table 1:** Baseline characteristics and laboratory findings of the studied population (n=85).

| Variable |  | All Cases<br>(n=85) |
| --- | --- | --- |
| Age (years) | Median (IQR) | 55 (42 – 65) |
|  | Range | 21 – 85 |
| Sex n, (%) | Male | 48 (56.5%) |
|  | Female | 37 (43.5%) |
| Comorbidities<br>n, (%) | DM | 31 (36.5%) |
|  | HTN | 34 (40.0%) |
|  | COPD | 7 (8.2%) |
|  | IHD | 5 (5.9%) |
|  | CLD | 3 (3.5%) |
|  | CKD | 15 (17.6%) |
| Symptoms n,<br>(%) | Cough | 1 (1.2%) |
|  | Cough and dyspnea | 2 (2.4%) |
|  | diarrhea | 1 (1.2%) |
|  | Dyspnea | 44 (51.8%) |
|  | fever | 34 (40.0%) |
|  | Fever and Cough | 3 (3.5%) |
| ICU<br>admission n,<br>(%) | Positive | 33 (38.8%) |
|  | Negative | 52 (61.2%) |
| Severity n,<br>(%) | Non-severe | 46 (54.1%) |
|  | severe | 39 (45.9%) |
| Laboratory<br>findings<br>Median (IQR) | Hemoglobin gm/dl | 12.6 (10.2 – 14.2) |
| | Platelets $\times 10^3/\text{cmm}$ | 227 (171.5 – 273.5) |
| | Total leukocytic count<br>$\times 10^3/\text{cmm}$ | 6.9 (4.35 – 10.55) |
| | Lymphocytes $\times 10^3/\text{cmm}$ | 1.34 (0.7 – 2.2) |
| | Neutrophils $\times 10^3/\text{cmm}$ | 4.28 (2.39 – 7.86) |
|  | Albumin (g/dl) | 3.5 (3 – 4.2) |
|  | CRP (mg/L) | 32 (9.5 – 132.5) |
|  | IL- 6 (pg/ml) | 55 (25 – 120) |
|  | L/CRP | 348.83 (99.78 – 1545.32) |
|  | CRP/alb | 0.94 (0.28 – 3.87) |
|  | L/IL-6 | 18.97 (8.25 – 51.71) |
| Hospital stay<br>(days) | Median (IQR) | 10 (6 – 17) |
|  | Range | (2 – 30) |
| Fate | Discharged | 64 (75.3%) |
|  | Died | 21 (24.7%) |

CKD: Chronic kidney disease; CLD: Chronic liver disease; COPD: Chronic obstructive pulmonary disease; DM: Diabetes mellitus; HTN: Hypertension; ICU: Intensive care unit; IHD: Ischemic heart disease; IQR: interquartile range.

### Characteristics of COVID-19 patients according to disease severity and in-hospital mortality

**Table 2** compares the baseline clinical characteristics and laboratory findings according to the severity and in-hospital mortality of COVID-19 patients.

**Table 2:** Comparisons of baseline characteristics and laboratory findings according to COVID-19 severity and in-hospital mortality.

| Variable |  | Non- severe<br>(n=46) | Severe<br>(n=39) | p- value | Survivors<br>(n=64) | Non-survivors<br>(n=21) | p- value |
| --- | --- | --- | --- | --- | --- | --- | --- |
| Age<br>(years) | Median<br>(IQR) | 55.5 (41.5 – 63) | 55 (43 – 67) | 0.250 | 55.5 (42 – 64.75) | 55 (47 – 69.5) | 0.454 |
| Sex n, (%) | Male | 23 (50.0%) | 25 (64.1%) | 0.191 | 34 (53.1%) | 14 (66.7%) | 0.277 |
|  | Female | 23 (50.0%) | 14 (35.9%) |  | 30 (46.9%) | 7 (33.3%) |  |
| Comorbidities n,<br>(%) | DM | 13 (28.3%) | 18 (46.2%) | 0.088 | 19 (29.7%) | 12 (57.1%) | 0.023 |
|  | HTN | 15 (32.6%) | 19 (48.7%) | 0.131 | 22 (34.4%) | 12 (57.1%) | 0.065 |
|  | COPD | 2 (4.3%) | 5 (12.8%) | 0.157 | 3 (4.7%) | 4 (19.0%) | 0.038 |
|  | IHD | 0 (0.0%) | 5 (12.8%) | 0.012 | 3 (4.7%) | 2 (9.5%) | 0.414 |
|  | CLD | 2 (4.3%) | 1 (2.6%) | 0.657 | 3 (4.7%) | 0 (0.0%) | 0.312 |
|  | CKD | 4 (8.7%) | 11 (28.2%) | 0.019 | 10 (15.6%) | 5 (23.8%) | 0.393 |
| Laboratory<br>parameters<br>Median<br>(IQR) | Lymp<br>×10<sup>3</sup>/cmm | 1.49<br>(1 – 2.22) | 0.9<br>(0.5 – 2.2) | 0.028 | 1.4<br>(0.77 – 2.2) | 0.9<br>(0.55 – 1.96) | 0.178 |
|  | CRP (mg/L) | 12<br>(6.5 – 32) | 129<br>(50 – 152) | ≤ 0.001 | 18<br>(7.7 – 65.75) | 152<br>(57.8 – 152) | ≤ 0.001 |
|  | Albumin<br>(g/dl) | 4<br>(3.1 – 4.8) | 3.2<br>(2.9 – 3.6) | ≤ 0.001 | 3.75<br>(3.02 – 4.35) | 3<br>(2.75 – 3.6) | 0.001 |
|  | IL- 6<br>(pg/ml) | 32<br>(18 – 50) | 120<br>(80 – 220) | ≤ 0.001 | 45<br>(20 – 97.5) | 168<br>(110 – 275) | ≤ 0.001 |
|  | L/IL-6 | 40.83<br>(25.20 – 98.71) | 8.33<br>(4.20 – 14.54) | ≤ 0.001 | 29.55<br>(11.90 – 79) | 5.33<br>(3.34 – 14.55) | ≤ 0.001 |
|  | CRP/alb | 0.35<br>(0.15 – 0.64) | 3.87<br>(1.66 – 4.90) | ≤ 0.001 | 0.52<br>(0.20 – 1.70) | 4.3<br>(1.82 – 5.52) | ≤ 0.001 |
|  | L/CRP | 1211.96<br>(449.82 – 2500) | 105.26<br>(56.96 – 230.26) | ≤ 0.001 | 703.27<br>(186.18 – 2005.35) | 105.26<br>(43.56 – 212.5) | ≤ 0.001 |
| ICU admission<br>n, (%) | Positive | 9 (19.6%) | 24 (61.5%) | ≤ 0.001 | 17 (26.6%) | 16 (76.2%) | ≤ 0.001 |
|  | Negative | 37 (80.4%) | 15 (38.5%) |  | 47 (73.4%) | 5 (23.8%) |  |
| Hospital stay<br>(days) | Median<br>(IQR) | 10.5 (6.75 – 17.5) | 9 (5 – 14) | 0.112 | 9.5 (6 – 17) | 10 (7 – 16) | 0.759 |
| Fate | Discharged | 43 (67.2%) | 21 (32.8%) | ≤ 0.001 |  |  |  |
|  | Died | 3 (14.3%) | 18 (85.7%) |  |  |  |  |

*CKD: Chronic kidney disease; CLD: Chronic liver disease; COPD: Chronic obstructive pulmonary disease; DM: Diabetes mellitus; HTN: Hypertension; ICU: Intensive care unit; IHD: Ischemic heart disease; IQR: Interquartile range; Lymp: Lymphocytes. Statistical significance set at 0.05.*

Significant changes between the severity groups were observed when their median (IQR) values were compared. As opposed to the non-severe group, severe patients had significantly higher values of CRP (129 (IQR: 50 – 152) vs.12 (6.5 – 32) mg/L; P ≤ 0.001), IL-6 (120 (80 – 220) vs. 32 (18 – 50) pg/ml; P ≤ 0.001) and CRP/alb (3.87 (1.66 – 4.90) vs. 0.35 (0.15 – 0.64); P ≤ 0.001). And, significantly lower values of L/CRP (105.26 (56.96 – 230.26) vs. 1211.96 (449.82 – 2500); P ≤ 0.001), L/IL-6 (8.33 (4.20 – 14.54) vs. 40.83 (25.20 – 98.71); P ≤ 0.001), lymphocyte count (0.9 (0.5 – 2.2) vs.1.49 (1 – 2.22) ×103/µL; P=0.028) and albumin (3.2 (2.9 – 3.6) vs. 4 (3.1 – 4.8) g/dl; P ≤ 0.001).

Regarding in-hospital mortality of COVID-19, significant differences between survivors and non-survivors were observed. CRP (152 (57.8 – 152) vs.18 (7.7 – 65.75) mg/L;P ≤ 0.001), IL-6 (168 (110 – 275) vs. 45 (20 – 97.5) pg/ml; P ≤ 0.001), and CRP/alb (4.3 (1.82 – 5.52) vs. 0.52 (0.20 – 1.70); P ≤ 0.001) showed significantly higher values in non-survivors compared to survivors. In contrast, L/CRP (105.26 (43.56 – 212.5) vs. 703.27 (186.18 – 2005.35); P ≤ 0.001), L/IL-6 (5.33 (3.34 – 14.55) vs. 29.55 (11.90 – 79); P ≤ 0.001) and albumin (3 (2.75 – 3.6) vs. 3.75 (3.02 – 4.35) g/dl; P= 0.001) were significantly lower. But, the change in lymphocyte count according to in-hospital mortality was not significant (0.9 (0.55 – 1.96) vs.1.4 (0.77 – 2.2) ×103/µL; P=0.178).

### Predictive performance evaluation

**Table 3:** shows diagnostic validity tests and ROC curve analysis used to assess each biomarker’s overall prognostic performance on its own, identifying those that performed well. Biomarkers with the best diagnostic efficacies (least false and highest true results) were then combined in a multi-ROC curve to maximize the AUC and improve predictive accuracy.

**Table 3:** Diagnostic validity and receiver operating curve (ROC) analysis for the ability of CRP, IL-6 and their derived indices to distinguish COVID-19 infected patients according to severity and mortality of the disease

| Indicators | Cut off | AUC | Specificity % | Sensitivity % | NPV % | PPV % | Accuracy % |
| --- | --- | --- | --- | --- | --- | --- | --- |
| **Severity** |  |  |  |  |  |  |  |
| CRP | 48 | 0.861 | 84.8 | 79.5 | 83.0 | 81.6 | 82.4 |
| IL- 6 | 50 | 0.873 | 78.3 | 87.2 | 87.8 | 77.3 | 82.4 |
| L/IL-6 | 11.66 | 0.883 | 95.7 | 71.8 | 80.0 | 93.3 | 84.7 |
| CRP/alb | 1.65 | 0.878 | 91.3 | 76.9 | 82.4 | 88.2 | 84.7 |
| L/CRP | 260.86 | 0.873 | 89.1 | 79.5 | 83.7 | 86.1 | 84.7 |
| CRP/alb at 1.65 + L/CRP at:2300 | -- | 0.922 | 91.3 | 100.0 | 100.0 | 90.7 | 95.3 |
| **In- hospital mortality** |  |  |  |  |  |  |  |
| CRP | 151 | 0.801 | 92.2 | 57.1 | 86.8 | 70.6 | 83.5 |
| IL- 6 | 120 | 0.888 | 92.2 | 71.4 | 90.8 | 75.0 | 87.1 |
| L/IL-6 | 5.40 | 0.866 | 95.3 | 57.1 | 87.1 | 80.0 | 85.9 |
| CRP/alb | 4.21 | 0.812 | 90.6 | 57.1 | 86.6 | 66.7 | 82.4 |
| L/CRP | 129.03 | 0.787 | 79.7 | 61.9 | 86.4 | 50.0 | 75.3 |
| IL-6 at 120 pg/ml + L/IL-6 at:8 | -- | 0.946 | 100.0 | 85.7 | 95.5 | 100.0 | 96.5 |

AUC: area under curve; CRP/alb = CRP (mg/dl)/albumin (g/dl); CRP: C-reactive protein in mg/L; IL-6: interleukin-6 in pg/mL; L/CRP = lymphocyte (number/ μl)/CRP (mg/dl); L/IL-6 = lymphocyte (number/ μl)/IL-6 (pg/ml); NPV: negative predictive value; PPV: positive predictive value.

Regarding COVID-19 severity, while CRP alone at a cut-off of 48 mg/L offered 0.861 AUC and 82.4% diagnostic accuracy, its derived indices, CRP/alb, and L/CRP, exhibited the best predictive performance compared to other biomarkers. At the cut-off point of 1.65 for CRP/alb and 260.86 for L/CRP, the AUC was 0.878 and 0.873, respectively, and the diagnostic accuracy was 84.7% for both. And when CRP/alb at a cut-off point of 1.65 was combined with L/CRP at a cut-off point of 2300 in a multi-ROC analysis, COVID-19 severity prediction accuracy increased to 95.3% with an AUC of 0.922. **Figure 1**

**Figure 1:**
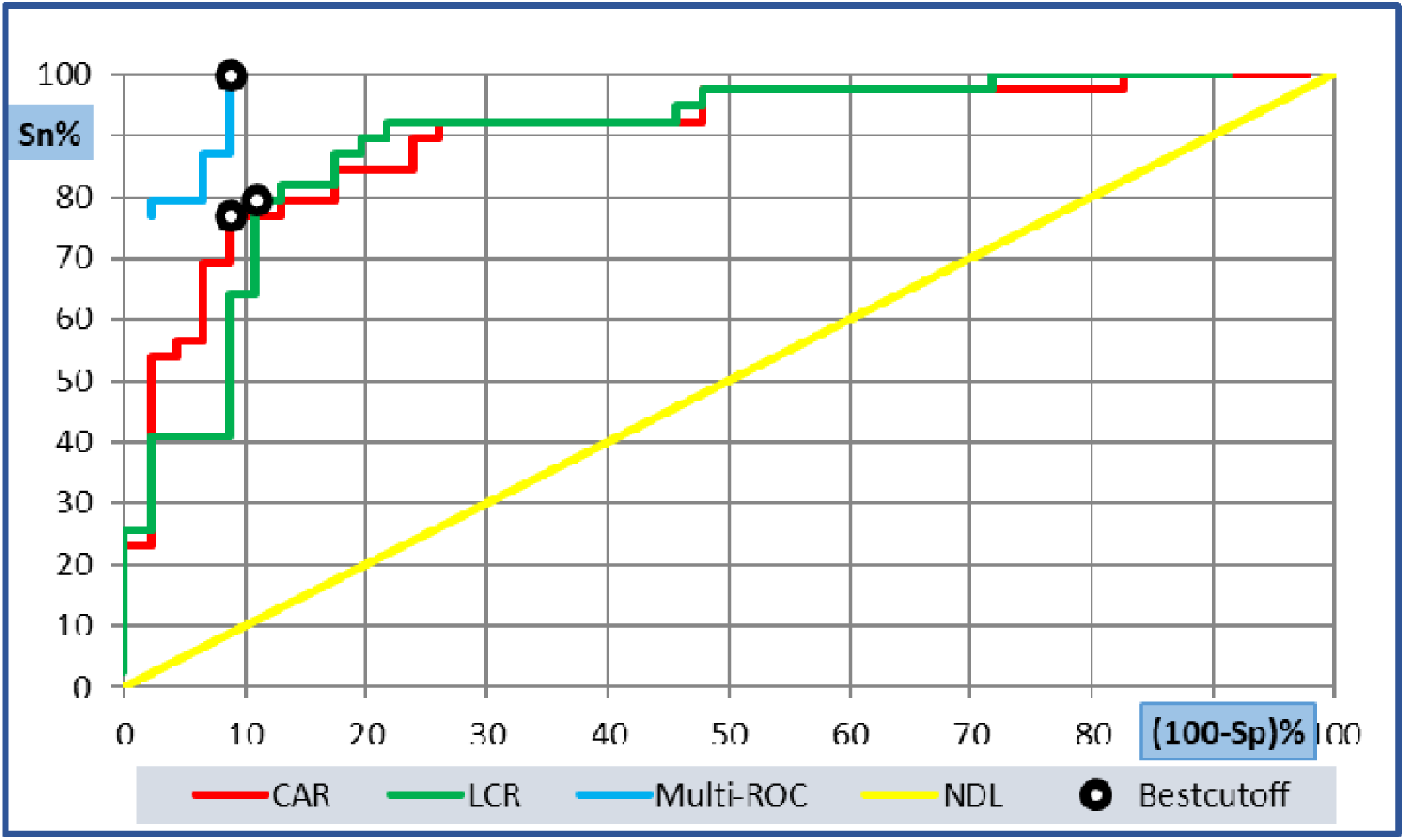
Diagnostic performance of CRP/alb (AUC:0.878; 95%CI: 0.805-0.952); L/CRP (AUC: 0.873; 95%CI: 0.798-0.948) and their combination (AUC: 0.922) for discriminating severe COVID-19 patients from those non-severe.

Concerning COVID-19 in-hospital mortality, the baseline IL-6 level was the most highly predictive biomarker with 0.88 AUC at a cut-off point of 120 pg/ml and diagnostic accuracy of 87.1%. Comparable to IL-6 was L/IL-6 with 0.86 AUC at a cut-off point of 5.40 and a diagnostic accuracy of 85.9%. Adding IL-6 at a cut-off point of 120 pg/ml to L/IL-6 at a cut-off point of 8 has improved the overall in-hospital mortality prediction ability and increased the AUC to 0.946 and the diagnostic accuracy to 96.5%. **Figure 2**

**Figure 2:**
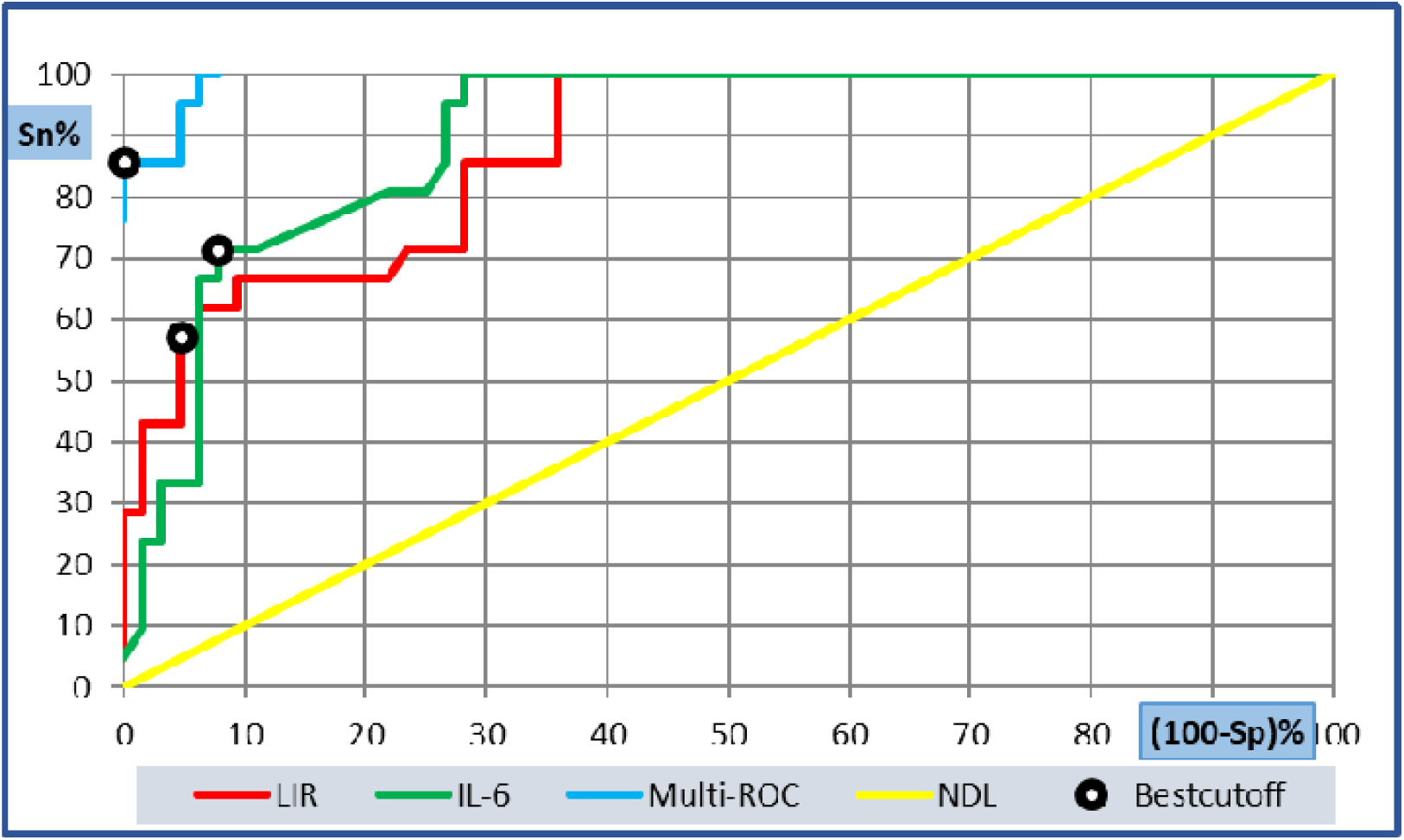
Diagnostic performance of L/IL-6 (AUC: 0.866; 95CI: 0.762-0.970); IL-6(AUC: 0.888; 95CI: 0.819-0.957), and their combination (AUC: 0.946) for discriminating survivors from non-survivors in COVID-19.

### Risk factors assessment

Logistic regression analysis further evaluated the association between IL-6, CRP, and their derived indices with COVID-19 severity and in-hospital mortality. As to COVID-19 in-hospital mortality, the multivariate analysis showed that the odds ratios (ORs) of all studied biomarkers were not significantly different (p > 0.05). And only IL-6 was an independent risk that was significantly positively associated with severe disease development (OR: 1.033; 95% CI: 1.002 - 1.066; P =0.037). **Table 4** demonstrates logistic regression analysis for possible predictors of COVID-19 severity and in-hospital mortality.

**Table 4:** Predictors of COVID-19 progression

| Indicator | Uni-variate |  |  |  | Multi-variate |  |  |  |
| --- | --- | --- | --- | --- | --- | --- | --- | --- |
|  | P-value | Odds ratio | 95% C.I. for OR |  | P-value | Odds ratio | 95% C.I. for OR |  |
|  |  |  | Lower | Upper |  |  | Lower | Upper |
| Mortality |  |  |  |  |  |  |  |  |
| L/IL-6 | **0.001** | 0.872 | 0.803 | 0.947 | 0.374 | 0.92 | 0.766 | 1.105 |
| CRP/alb | **≤ 0.001** | 1.921 | 1.431 | 2.579 | 0.439 | 2.602 | 0.231 | 29.268 |
| L/CRP | **0.065** | 0.999 | 0.999 | 1 | 0.164 | 1 | 1 | 1.001 |
| CRP (mg/L) | **≤ 0.001** | 1.021 | 1.011 | 1.031 | 0.758 | 0.988 | 0.917 | 1.065 |
| IL– 6 (pg/ml) | **≤ 0.001** | 1.019 | 1.01 | 1.028 | 0.152 | 1.013 | 0.995 | 1.032 |
| Severity |  |  |  |  |  |  |  |  |
| L/IL-6 | **0.001** | 0.945 | 0.914 | 0.976 | 0.652 | 1.006 | 0.981 | 1.032 |
| CRP/alb | **≤ 0.001** | 2.787 | 1.821 | 4.264 | 0.371 | 7.401 | 0.092 | 593.008 |
| L/CRP | **≤ 0.001** | 0.997 | 0.996 | 0.999 | 0.285 | 0.999 | 0.996 | 1.001 |
| CRP (mg/L) | **≤ 0.001** | 1.03 | 1.018 | 1.043 | 0.495 | 0.956 | 0.84 | 1.088 |
| IL– 6 (pg/ml) | **≤ 0.001** | 1.032 | 1.018 | 1.047 | **0.037** | 1.033 | 1.002 | 1.066 |

CI: Confidence interval

## Discussion

During the current pandemic of COVID-19, since immune dysfunction, hyper-inflammation, and hyper-cytokinaemia have been closely linked to the rapid progression of the disease,^**6**^ continuous finding of sensitive inflammatory biomarkers for timely and effective identification of disease progression is necessary.^**7**^

CRP, an inflammatory molecule that plays a vital role in host resistance to infections, was highly linked to acute lung injury and unfavorable outcomes in COVID-19 patients. As a result, detecting CRP levels is quite useful in determining the severity of COVID-19 patients.^**8**^

In our study, CRP increased significantly with COVID-19 severity and mortality. Similarly, several studies have reported significantly higher CRP values in the more severe patients and those who died from the disease.^**8-10**^ On the contrary, Zhang Y. revealed no significant change in CRP levels in patients after being admitted to ICU.^**11**^ Also, Ferrari and colleagues, based on statistical analysis, stated that CRP might not be very useful in discriminating between patients with or without COVID-19.^**12**^

IL-6 is a multifunctional cytokine that regulates immunological and inflammatory responses. It plays a key role in the development and progression of coronavirus pneumonia. In COVID-19 patients with the hyper-inflammatory syndrome, circulating IL-6 concentrations have been linked to disease severity, the incidence of acute lung damage, and mechanical ventilation need, suggesting that measuring IL-6 levels can help guide therapy decisions.^**13**^

In our study, IL-6 levels were significantly higher in severe patients and non-survivors. Per our results, Sun and co-workers reported significantly higher IL-6 values in the more severe patients.^**13**^ Also, Coomes and Haghbayan have reported similar results in their systemic review.^**14**^ Furthermore, several studies have reported the distinct clinical significance of IL-6 at varying cut-off values. Gao et al. studied a cohort of 43 cases and reported that combining IL-6 at a cut-off value of 24.3 pg/ml with the D-Dimer helped in the early detection of severity progression.^**15**^ In another cohort by Giofoni et al., serum IL-6 at a cut-off value of 25 pg/ml represented an independent risk of COVID-19 progression.^**16**^ In a cohort study of 40 patients in Munich, COVID-19 patients’ need for mechanical ventilation increased 22 times with the elevated IL-6 levels (> 80 pg/ml).^**17**^

In a meta-analysis, Coomes and Haghbayan discovered statistical heterogeneity in IL6 levels in subjects with varying degrees of disease severity. They attributed this to inconsistencies of patients’ characteristics in each trial and discrepancies in the timing of IL6 testing and immunomodulatory medicines received.^**14**^

Significant hypo-albuminemia and lymphopenia were discovered in severe patients in the current study; however, only significant hypo-albuminemia and not lymphopenia was detected in non-survivors. Returning to the fact that albumin levels drop in inflammatory conditions, hypoalbuminemia has been noted in COVID-19 patients with severe hyperinflammation.^**18**^

In several studies,^**3**, **19**^ COVID-19 severity and progression have been linked to lymphopenia, a reliable and accurate prediction. According to a meta-analysis study, the more serious the condition, the fewer lymphocytes there were ^**20**^, which could be attributed to the disease’s effect on T cell subsets, resulting in immunological dysfunction.^**21**,**22**^ Several studies have emphasized that when the lymphocyte count is less than 1.5 10^3^/µL, lymphopenia should be addressed.^**9**^ Using an inflammatory biomarker alone to evaluate patients infected with COVID-19 could be influenced by several factors. As a result, the generated indices from the most important inflammatory biomarkers may more accurately and thoroughly indicate immunological dysregulation.^**7**, **23**^

For numerous malignancies, including colon and stomach carcinomas, the L/CRP has been employed as a prognostic marker because it’s a good predictor of the complicated host-tumor immunological interactions that lead to the systemic inflammatory process implicated in the etiology of these carcinomas.^**24-26**^ COVID-19 also triggers an inflammatory reaction throughout the body; therefore, the L/CRP could be a useful prognostic biomarker for disease severity and outcome. On exploring the role of L/CRP in the prognosis of our studied COVID-19 patients, we found that it declined significantly with the increase in disease severity and mortality. By our results, Ullah and co-workers have found that a low L/CRP was a good predictor of complications and mortality in COVID-19 patients. ^**24**^

High CRP/alb was previously linked to mortality in critically ill patients of various conditions.^**27**^ In our study, we found a significant elevation of the CRP/alb as the disease progressed. Similarly, El-Shabrawy and colleagues have found that the CRP/alb was a predictive factor of COVID-19 mortality.^**28**^

In our study, the L/IL-6 was much lower in severe COVID-19 patients and non-survivors. In the same context, because high IL-6 levels and low lymphocyte counts have been connected to the severity and in-hospital mortality of COVID-19, in a study by Yang and colleagues, the IL-6/lymphocyte ratio (IL-6/L, reversed) was described as a novel biomarker to conduct risk stratification in COVID-19 patients.^**7**^ Other studies have found that having a high IL-6/L is an independent risk factor for disease progression and a poor prognosis.^**29**, **30**^ The decrease in lymphocyte numbers caused by IL-6-mediated inhibition of T cell activation explained the high in COVID-19.^**31**^ Furthermore, in a study by Liu et al., T cell numbers were inversely associated with the studied cytokine levels, including IL-6.^**32**^ As a result, the balance of IL-6 and lymphocytes appeared to be crucial in the immune system’s homeostasis.

In the multi-ROC curve analysis, we integrated biomarkers that exhibited the best predictive performance for COVID-19 progression to achieve the maximum overall test efficacy and improve the predictive ability. The combination of CRP/alb and L/CRP improved disease severity prediction accuracy. Similarly, combining IL-6 with the L/IL-6 enhanced in-hospital mortality prediction accuracy. Finally, only IL-6 was positively correlated with disease severity, but none of the investigated biomarkers were shown to be an independent predictor of in-hospital mortality. This could be attributed to the small sample size and the disproportion between the study groups, and these were the study’s major limitations.

More research on a larger scale and for a longer period is needed to determine the predictive usefulness of the proposed inflammatory indices compared to the other existing biomarkers to slow the disease’s progression and put the epidemic to an end.

## Conclusions

The current study discovered that CRP and IL-6 and their derived immune-inflammatory indices, namely CRP/alb, L/CRP, and L/IL-6, were good predictors of COVID-19 progression. Using these indicators in combinations rather than a single biomarker alone would improve the overall performance in disease progression prediction.

## Data Availability

Data will be available upon request.

## Abbreviations

CRP/alb: CRP/albumin
L/CRP: lymphocyte/CRP
L/IL-6: lymphocyte/IL-6

## Acknowledgement

None.

## Authors’ contribution

All authors contributed significantly to this work, whether it was in the conception, study design, data acquisition, analysis, and interpretation, or in the drafting, revising, or reviewing of the article, and they all provided final approval of the version to be published.

## Data availability statement

On reasonable request, the corresponding author will provide the datasets used and/or analyzed during the current work.

## Ethics

Ethical approval for the current study protocol was obtained from Ain Shams University Faculty of Medicine Research Ethics Committee (REC) FWA 00017585. All patient data were kept private and confidential. Solely utilized them for research purposes.

## Statement of informed consent

All procedures were explained to all participants or their first-degree relatives, with informed consent obtained from them.

## Consent for publication

Not applicable.

